# An analysis of mortality in Ontario using cremation data: Rise in cremations during the COVID-19 pandemic

**DOI:** 10.1101/2020.07.22.20159913

**Authors:** Gemma Postill, Regan Murray, Andrew S. Wilton, Richard A. Wells, Renee Sirbu, Mark J. Daley, Laura C Rosella

**Affiliations:** Department of Epidemiology and Biostatistics, Western University; Office of the Chief Coroner for Ontario, 25 Morton Shulman Drive, Toronto, ON, Canada; Public Health Agency of Canada; ICES, 2075 Bayview Ave, Toronto, ON, Canada; Dalla Lana School of Public Health, University of Toronto, 155 College Street, Suite 600, Toronto, M5T 3M7, ON, Canada; The Vector Institute for Artificial Intelligence, Toronto, ON, Canada; Institute for Better Health, Trillium Health Partners, Mississauga, ON, Canada

**Keywords:** Mortality, Cremation, Covid-19, SARS-CoV-2

## Abstract

**Background:** The impact of coronavirus disease 2019 (COVID-19) on mortality in Ontario is unknown. Cremations are performed for most deaths in Ontario and require coroner certification before the cremation can take place. Our objective was to provide timely analysis of deaths during the COVID-19 pandemic using cremation data.

**Methods:** We analyze cremation certificate data from January 1, 2017, to June 30, 2020, in Ontario. 2020 cremation records were compared to historical records from 2017-2019 by age, month, and place of death and COVID-19 status. A time series model was fit to quantify the deviation in cremation trends during the COVID-19 period.

**Results:** There have been 39 760 cremations in Ontario in 2020 with the highest number of seen in April (N = 7 527 cremations) when peak COVID-19 cases were seen. Over the study period, the proportion of cremations from deaths in hospitals decreased whereas cremations from long-term care and residences increased. In April there were 1 839 more cremations compared to historical averages over 2017-2019, representing a 32% increase. Time series modelling of cremations from January 2017 demonstrated that cremations in April and May 2020 exceeded the projections based on modelled estimates.

**Conclusion:** We demonstrate the utility of cremation data for providing timely mortality information during a public health emergency. Cremations were higher in the pandemic months compared to previous years, and there was a shift in deaths occurring in hospitals to long-term care and residences. These timely estimates of mortality are critical for understanding the impact of COVID-19.

## Introduction

The 2019 novel coronavirus (SARS-CoV-2) has given rise to the severe COVID-19 disease around the world. COVID-19 has had a significant impact on health systems and mortality globally. While the impact on hospitalizations and intensive care units is routinely tracked, mortality data is less consistent, given the variation in how mortality data are captured. Quantifying the effect of COVID-19 on all-cause mortality is a critical element needed to inform the policy response.

In Canada, the responsibility of death recording and reporting deaths lies with the provinces and territories, according to the Vital Statistics Act.^1^ There are several challenges with obtaining real-time mortality data given the need to verify the record and cause of death, which can take months or years. Although COVID-19 mortality has been reported in Ontario through public health databases, two of the number of deaths from other causes is not yet available as the routine verification process that results in the official Vital Statistics records can lag more than a year. Without all-cause mortality information, the data on COVID-19 specific mortality is difficult to contextualize. Further, there remain critical questions regarding the mortality during the pandemic from all causes given restrictions to medical care and other consequences of large-scale shutdowns. Several other countries have reported increases in morality during the COVID-19 period;^3-5^ however, the extent of this effect in Canada is unknown and required to inform both healthcare and public health responses.

In Ontario, in recent years, cremations are performed for about 70% of deaths.^6^ Before cremation can occur, a coroner’s certification is needed.^7^; as a result, cremation data are available in near-real-time and offers a consistent data source to examine these mortality trends in a timelier manner. Cremation data has been used in other countries to quantify the mortality trends during the COVID-19 pandemic. Our goal was to estimate the COVID-19 mortality from cremation data across age and place of death and to quantify the difference in those trends by comparing them to historical cremation records for 2017-2019 to generate the earliest analysis of mortality during the pandemic in Ontario.

## Methods

### Study Population and Data Collection

In Ontario, Canada, a Cremation Certificate must be provided by a coroner to authorize the cremation of a deceased person.^7^ A licensed crematorium operator cannot proceed without a Cremation Certificate. The law in Ontario requires that a coroner review the circumstances surrounding the death before cremation takes place. The review and authorization are documented on the certificate and kept on file at the crematorium.^1^ Since 2017, these records have been collected and stored electronically by the Office of the Chief Coroner for Ontario. This database contains names, dates of birth and death, location of death, and cause of death for every person cremated in the province since that time. Records submitted until July 27 with dates of death before July 2020 were included. All cremations were de-identified and maintained in an electronic database for the analysis. Records with a cause of death specified as ‘test’ or age at death greater than 120 were excluded.

The 2020 COVID-19 related deaths were isolated from the 2020 records by the presence of the terms COVID or novel coronavirus as the cause of death, antecedent cause, or other cause categories. These were the only terms found to describe COVID in the cremation records. Records that matched the above criteria but also contained the following phrases: test-results pending, possible, and negative, were excluded from the classification of death due to COVID-19.

### Statistical Analysis

The records were categorized according to the month of death and subcategorized by age and type of facility in which death occurred. Age in the file is a numerical field, which was converted into a categorical vector (0-44 years, 45-74 years, and 75+ years). The facility at the time of death is reported as either long-term care, hospital, residence, or other. The stratified monthly difference in 2020 cremations relative to the average number of cremations by month in 2017-2019 was calculated as both the absolute difference and percent increase.

The number of weekly cremations (Monday to Sunday) was determined for 2017, 2018, and 2019 to demonstrate the expected annual seasonality in the number of cremations. Their respective trends were smoothed using the Statsmodel Holt’s package; the default additive model was changed to an exponential model with a fixed smoothing slope (β = 0.2) and smoothing level (α = 0.6). Exponential smoothing was used as the data was demonstrated with the Augmented Dickey-Fuller Test to be non-stationary (ADF Test Statistic of -2.499 with a P-Value 0.116). The number of weekly cremations for 2020 was tabulated both with and without COVID-19 deaths to determine the variance in cremations with and without COVID-19.

Using the daily cremation counts from January 2017, a time series model was fit with FB Prophet, an automated additive regression model with adjustable parameters model the historical cremation trends and estimate the projected number of cremations that would be expected for 2020 if historical trends continued. The parameters used in the model were a logistic growth curve and a yearly seasonality component. The yearly seasonality component was estimated with a partial Fourier sum of order 10. The flexibility of the trend was increased by adjusting the change point parameter to 0.05, and the confidence interval was set to 95%. Outliers beyond a 99% confidence interval (N=191) were removed, and the data was re-modelled with the same methodology. The following 365 days (2020) were forecasted to determine the expected number of cremations for 2020. The actual values for weekly cremations were overlaid on the 95% confidence interval of the time series to compare. All analysis was done using Python 3.7.0.

## Results

Of the 277 552 cremation records analyzed, 240 had missing data for cause of death, 4 419 records had missing age at death, and 180 had a missing facility of death. Overall, there were 41 280 cremations in the year 2020, and Table 1 shows that the distributions of these cremations shifted to greater proportions seen in deaths occurring in Long Term Care (LTC) and personal residences. Of the total cremations in 2020, more than 50% took place in hospital before April whereas the proportion of cremations from deaths occurring in hospital fell to 42%, 45% and 46% in April, May and June respectively. Over the same period, an increase in the number and distribution of cremations occurring in long term care and personal residences were seen. In the peak month (April) there were 1 974 cremations from deaths occurring in long term care, representing 25% of all cremations. Cremation from deaths in personal residence rose steadily each month with the highest numbers seen in April (2 050 deaths, 26% of April cremations) and May (2 054; 29% of May cremations). Cremations by month and age are shown in Table 2. 26, 892 (68%) took place from March to June, corresponding to the pandemic period in Ontario. The highest number of cremations (N=7 527) was observed in the month of April, which was also when the most COVID-19 cases were confirmed (N=14 435 cases). The majority of the April cremations (63%, N=4 754) occurred in the 75 and older age group; this group demonstrated the highest increase compared to the 2017-19 baseline (37.7%). The number of cremations in both January and February showed small increases over the 2017-2019 average for each age of death category (2.3% and 4.5% respectively). In contrast, the numbers of cremations in March, April, and May differed substantially from the 2017-2019 historical averages with an 8.4% increase in March, a 32% increase in April and a 20% increase in May, and a 10% increase in June of 2020 (Table 2). In Ontario, more than 70% of deaths proceed to cremation, with higher percentages in those aged 55 and older and smaller numbers in the younger ages (<19 years) (Supplement Figure 1).

**Table 1.** Total number of and percentage of cremations by month and facility type in 2020

| Month of Death in 2020 | Facility of Death | Number of Cremations in 2020 | Percent of Cremations in 2020 | Average Number Cremations 2017-2019 | Percent of 2017-2019 Cremations |
| --- | --- | --- | --- | --- | --- |
| January | Hospital | 3 777 | 53.5 | 3 731 | 53.9 |
|  | Long-term Care | 1 298 | 18.4 | 1 328 | 19.2 |
|  | Residence | 1 436 | 20.3 | 1 369 | 19.8 |
|  | Other | 553 | 7.8 | 491 | 7.1 |
|  | **Total** | **7 064** | **100.0** | **6 919** | **100.0** |
| February | Hospital | 3 362 | 53.7 | 3 218 | 53.4 |
|  | Long-term Care | 1 102 | 17.6 | 1 171 | 19.4 |
|  | Residence | 1 320 | 21.1 | 1 220 | 20.2 |
|  | Other | 475 | 7.6 | 420 | 7.0 |
|  | **Total** | **6 259** | **100.0** | **6 029** | **100.0** |
| March | Hospital | 3 602 | 51.9 | 3 475 | 54.3 |
|  | Long-term Care | 1 254 | 18.1 | 1 170 | 18.3 |
|  | Residence | 1 589 | 22.9 | 1 286 | 20.1 |
|  | Other | 496 | 7.1 | 466 | 7.3 |
|  | **Total** | **6 941** | **100.0** | **6 397** | **100.0** |
| April | Hospital | 3 269 | 41.9 | 3 215 | 54.3 |
|  | Long-term Care | 1 974 | 25.3 | 1 044 | 17.7 |
|  | Residence | 2 050 | 26.3 | 1 214 | 20.5 |
|  | Other | 513 | 6.6 | 443 | 7.5 |
|  | **Total** | **7 806** | **100.0** | **5 916** | **100.0** |
| May | Hospital | 3 163 | 44.7 | 3 231 | 54.6 |
|  | Long-term Care | 1 384 | 19.6 | 984 | 16.6 |
|  | Residence | 2 054 | 29.0 | 1 223 | 20.7 |
|  | Other | 476 | 6.7 | 481 | 8.1 |
|  | **Total** | **7 077** | **100.0** | **5 920** | **100.0** |
| June | Hospital | 2 890 | 47.1 | 3 037 | 54.2 |
|  | Long-term Care | 901 | 14.7 | 897 | 16.0 |
|  | Residence | 1 860 | 30.3 | 1 196 | 21.4 |
|  | Other | 482 | 7.9 | 469 | 8.4 |
|  | **Total** | **6 133** | **100.0** | **5 600** | **100.0** |
| All 2020<br>(January to June) | Hospital | 20 063 | 48.6 | 19 907 | 54.1 |
|  | Long-term Care | 7 913 | 19.2 | 6 595 | 17.9 |
|  | Residence | 10 309 | 25.0 | 7 509 | 20.4 |
|  | Other | 2 995 | 7.3 | 2 771 | 7.5 |
|  | **Total** | **41 280** | **100.0** | **36 781** | **100.0** |

**Table 2.**
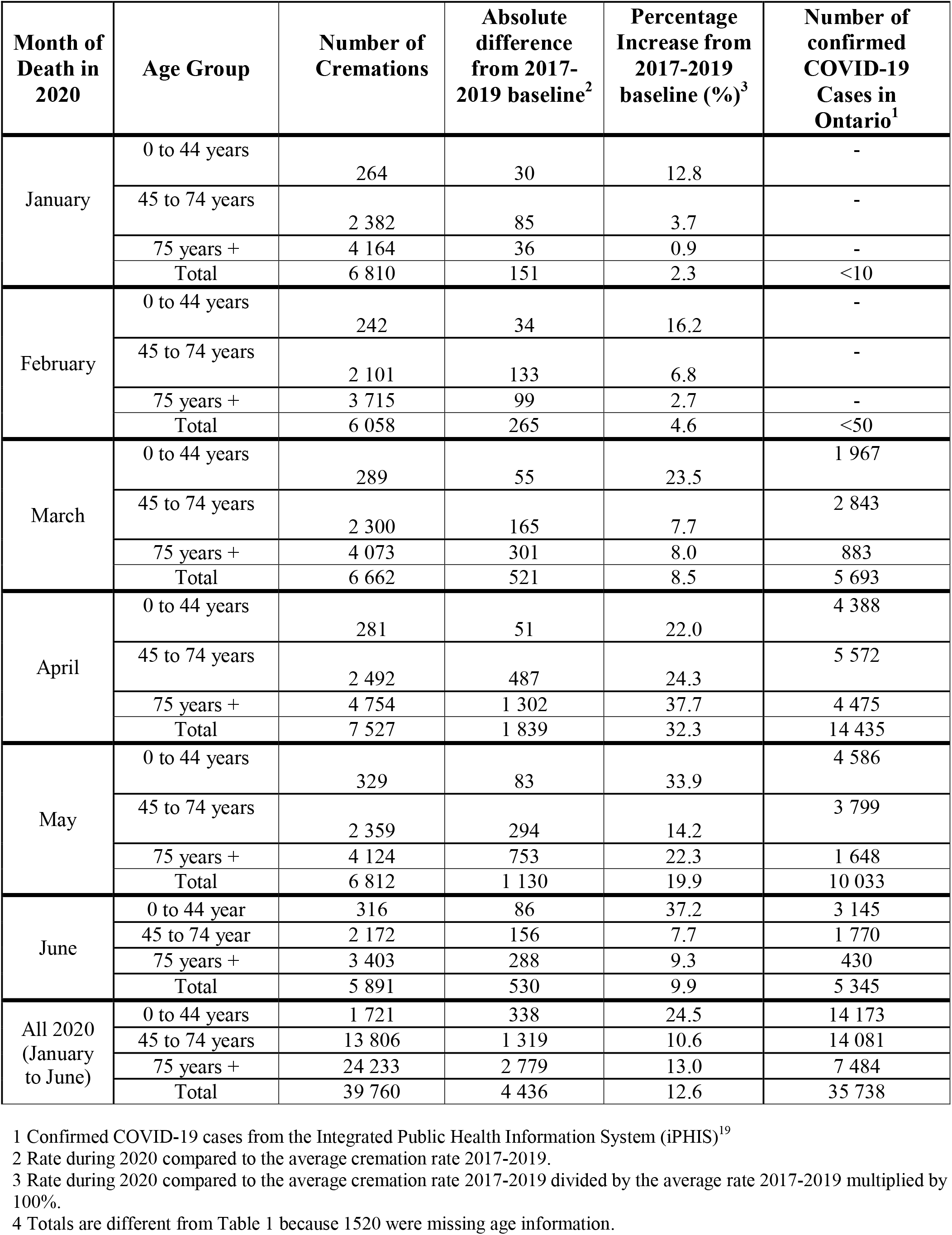
Number of monthly cremations in 2020 by age group compared to the average of 2017-2019

The distribution of facility of death for individuals cremated in Ontario also shifted during 2020. The differences in all-cause cremation numbers vary by the facility of death and by month such that the greatest differences were seen in long-term care homes in the month of April (89% increase compared to April in the 2017-19 period), which was also the month with the highest numbers of confirmed COVID-19 deaths reported by public health (N=1 399). Notably, the increase in cremations for deaths occurring at home was also substantially higher in March (23%), and April (68%) (Figure 1), and most of the cremations in April were among deaths in long-term care settings (Supplement Figure 2). A similar number of cremations from deaths occurring in hospitals during 2020 was seen compared to historical averages; however, the distribution of deaths occurring in long-term care and in residences was higher in the 2020 period compared to 2017-2019 (Supplement Figure 2).

**Figure 1:**
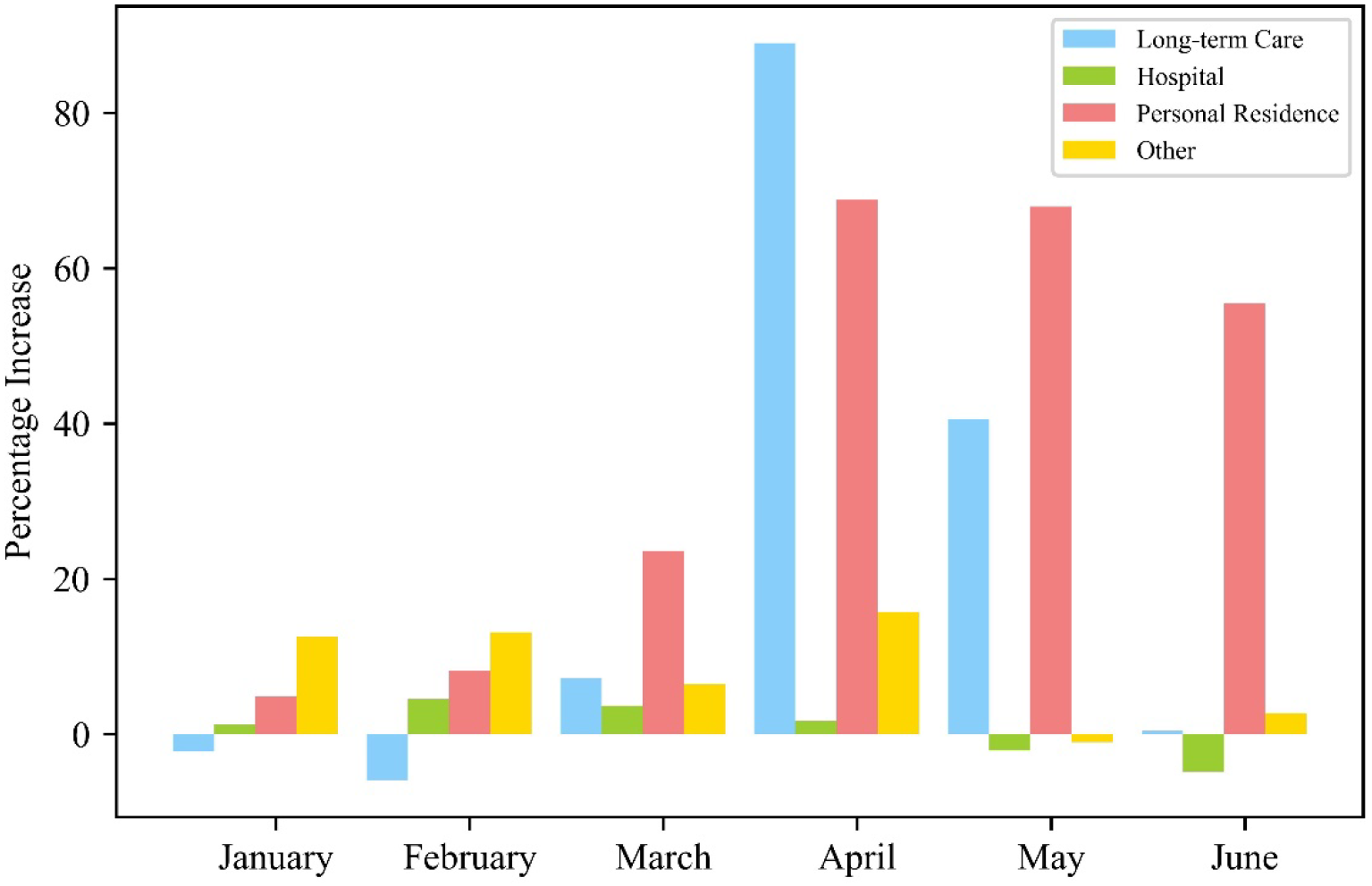
Percentage increase in the number of monthly cremations in 2020 by facility at time of death compared to the average of 2017-2019.

A time-series methodology was used and demonstrated that the number of daily cremations for March, April, and May 2020 exceeded the predicted increase of our time series model with 95% CI bounds, as illustrated in Figure 2. The model demonstrated that there had been an overall increase in the number of cremations in Ontario beyond what was predicted by the model, with 1,143 more cremations observed that projected for April and 403 more in May. (Supplement Figure 2)

**Figure 2:**
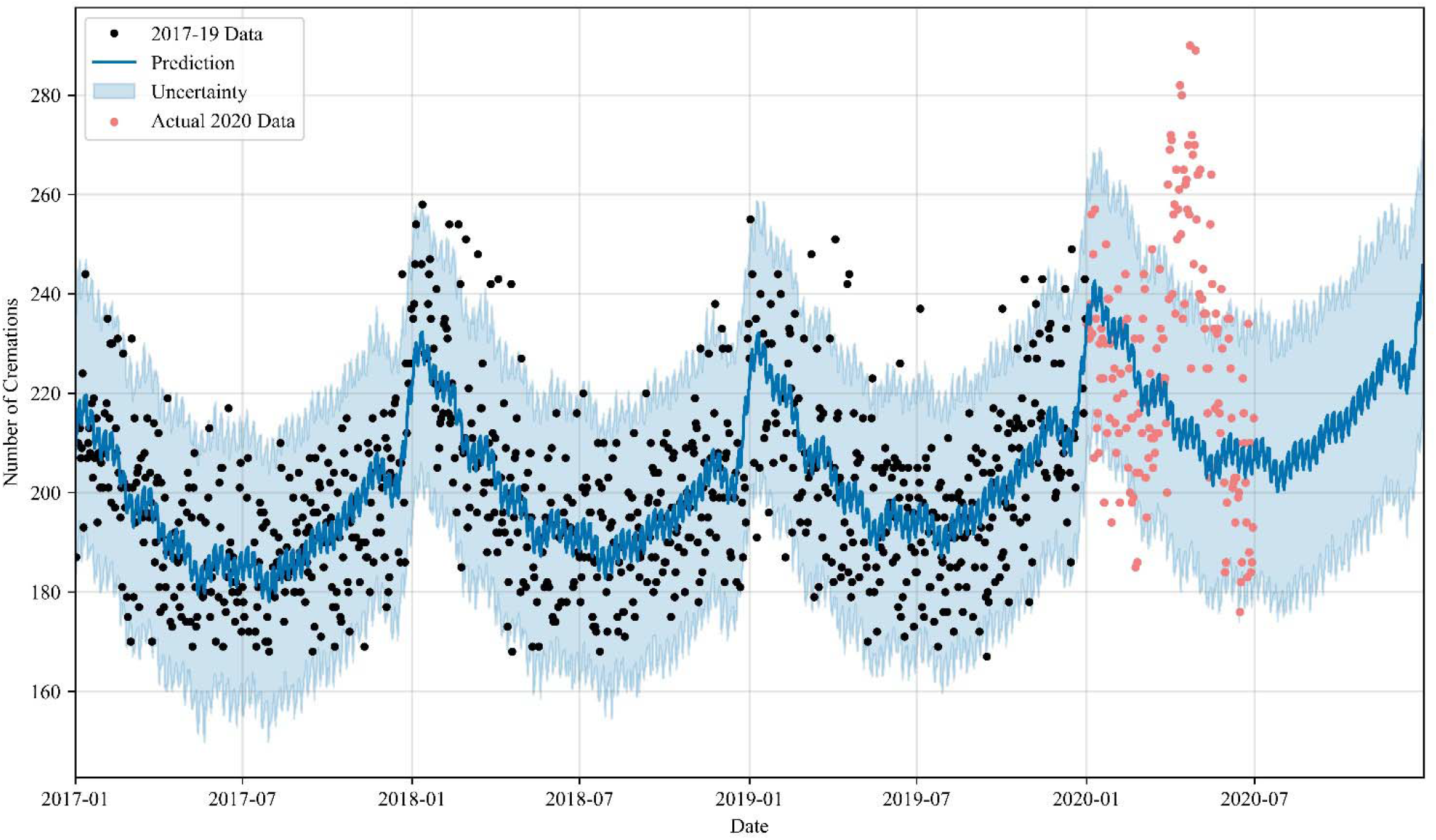
Weekly mortality rate from January 1, 2020 to June 30, 2020. The time series model is represented by the blue solid line and 95% confidence intervals in the shaded blue band.

Weekly cremation trends separating deaths related to COVID-19 are shown in Figure 3. COVID-19 deaths are elevated and account for a substantial proportion, but not all, the increase seen in April and May. The number of COVID-19 cremations by month are 50 in March, 867 in April, 604 in May, and 129 in June. Even excluding reported COVID-19 cremations, the monthly trends for 2020 are substantially different from 2017, 2018, and 2019 exponentially smoothed trends shown in Figure 3.

**Figure 3:**
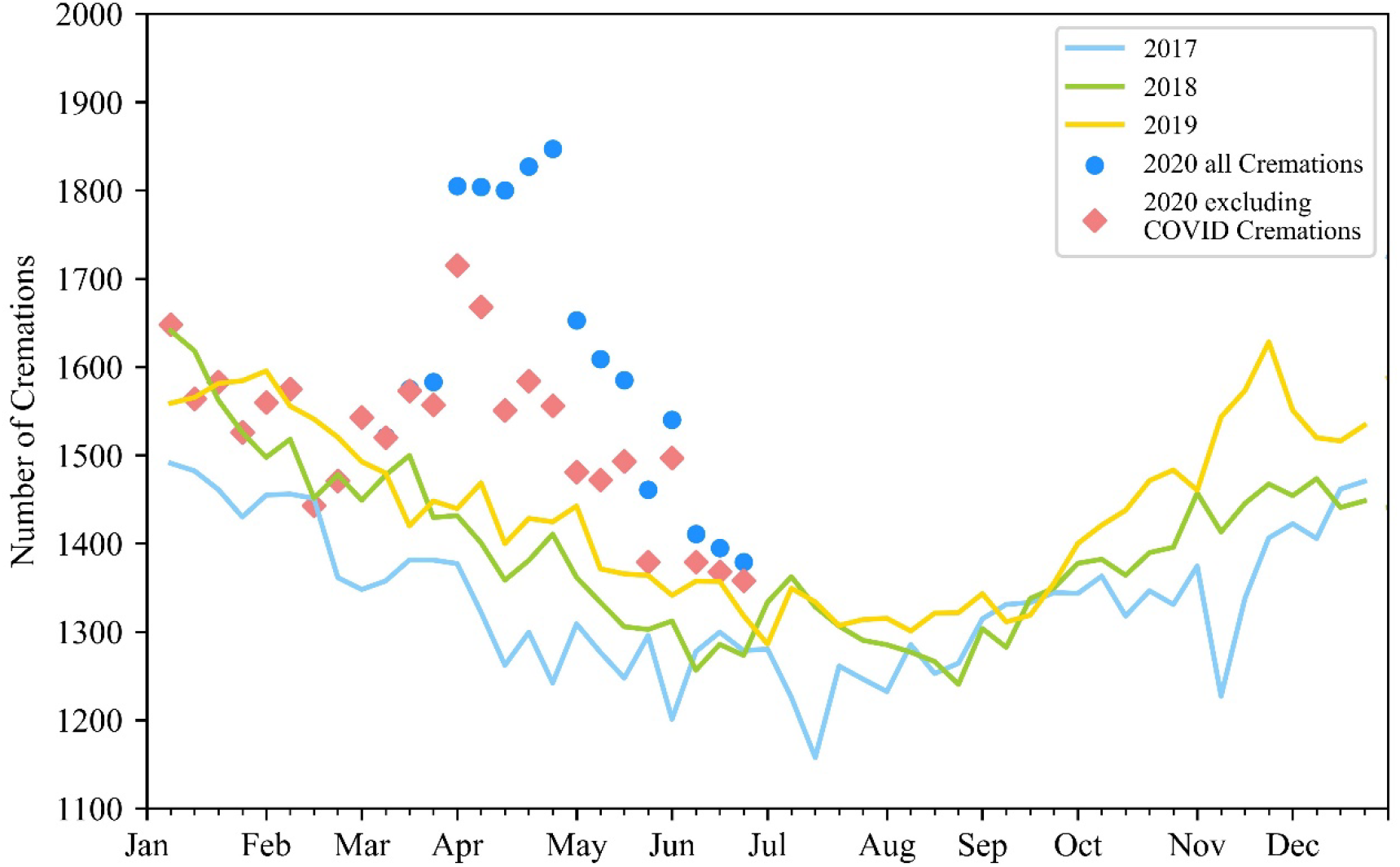
Smoothed trend of the number of cremations for 2017-2019 overlay with COVID-19 and non-COVID19 cremations for 2020 using an exponential model with a fixed smoothing slope (β = 0.2) and smoothing level (α = 0.6).

Owing to the lack of overall mortality data currently available in Ontario, we are unable to directly measure the cremation rate in the province during the pandemic period. However, for the period April 15 - May 31, 2020, mortality data from long-term care homes have been tracked by the Office of the Chief Coroner in real-time. We used these data along with mortality data for Ontario long-term care homes for 2017, 2018, and the first three quarters of 2019, to calculate the cremation rate in this population (Supplement Table 1). The proportion of deaths occurring in long-term homes during the peak period of the COVID-19 in Ontario has not increased in comparison to previous years.

## Discussion

This is the first study to provide estimates of mortality trends for Ontario during the COVID-19 pandemic and to demonstrate the utility of cremation data in monitoring mortality. Our results identify a significant increase in the number of cremations during March, April and May of 2020, which coincide with COVID-19 activity in Ontario. While COVID-19 deaths are routinely reported, monitoring all-cause mortality is an important metric, critical to contextualizing these numbers and understanding the full impact of COVID-19 in the population. The strengths of the study are the inclusion of all cremations up to the end of June 2020 and the inclusion of all-cause mortality in addition to COVID-19 deaths.

The results demonstrate an apparent increase in all-cause cremations during the months where the number of COVID-19 cases was highest in Ontario. The observed increases far exceed those predicted according to the time series model. This supports findings from many other regions in the world, reporting excess mortality during the COVID-19 pandemic.^3 4^ As found in other studies, not all of the mortality increase was due to reported COVID-19 deaths alone. Estimates from other jurisdictions attribute 50-80% of the rise due to reported COVID-19 directly, which was consistent with what was observed in the cremation data. There are several reasons why non-COVID-19 deaths may also increase, including delay in emergency^9^ and routine medical care for cardiovascular^10^ and other conditions^11^ as well as other factors such as fears of dying in isolation in hospital.^12^ Provisional vital Vital Statistics data had recently been made available for individual provinces (not Ontario), and in those data excess mortality was observed in British Columbia, Quebec and Alberta

We noted a substantial increase in the number of cremated deaths occurring from deaths in long-term care, which is consistent with the high burden of COVID-19 in this population.^14 15^ We also noted an increase in the number and proportion of deaths occurring in personal residence compared to the portion of deaths seen in hospitals in previous years. This indicates that the increase in mortality was not exclusively seen in long-term care but also included an increase in deaths occurring in personal residences.

A portion of the observed increase in late March and early April 2020 may be due to under-recognition of COVID-19 mortality early in the pandemic. Furthermore, there could be an under-recognized number of COVID-19 deaths throughout the pandemic period due to a lack of testing or false-negative test results.^16^ The variation in April and May 2020 could be due to several factors, including a shift toward cremation over burial in response to restrictions on travel, social gatherings, finances, or an increase in all-cause mortality secondary to COVID-19 pandemic response and wide-spread societal, health, and social service changes. As Ontario maintained social gathering restrictions and a state of emergency in May, the spike in cremations in April and subsequent decline in May supports an increase in all-cause mortality due to other factors such as delayed medical care, the loss of employment/income,^17^ isolation, and/or under-recognition of COVID-19 mortality.

There are some important limitations to this analysis that should be considered. First, specific cause-of-death data were not included because of the nature of the data structure (open text fields) and the heterogeneity in the classification of cause of death as immediate and antecedent, which prevents rapid analysis. Additional cremation records may be recorded for the 2020 time period, but this is not expected to change the findings reported in this study. As discussed, while cremation data represent the majority of deaths in Ontario, they do not represent all deaths. Certain segments of the population may be more or less likely to be cremated. This would be an issue in the comparison if the number of deaths during the pandemic period was affected disproportionately in a group with an intrinsically higher or lower rate of cremation (e.g. age group, burial traditions, geographic regions). Therefore, we acknowledge that at least some of the changes observed may be due to differences in preferences for cremation during the pandemic. However, we do not anticipate this to be the main reason for the increase given the already very high numbers of cremations before the pandemic (>70%), the magnitude of excess cremations (>30% during the peak month of COVID-19 activity) and the congruence of the increase and subsequent decreases with the confirmed COVID-19 activity. Using long-term care data we further demonstrate that the cremation rate is not meaningfully different than in previous years, making it very unlikely for changing in cremation practices alone to explain the excess mortality seen. Furthermore, we note that there was no provincial guidance against embalming during the outbreak,^18^ which would limit the likelihood of burial policies influencing the results. The relationship between cremation numbers and overall mortality will be verifiable when vital statistics data for this period becomes available.

In summary, we observed an increase in the number of cremations in the Province of Ontario during the COVID19 pandemic. The number of cremations was significantly higher during the peak pandemic period in Ontario compared to previous months and historical averages. This study documents the first changes in mortality during the COVID-19 pandemic in Ontario, and these data will continue to be essential to inform the ongoing COVID-19 pandemic response. More studies are required to confirm further the nature of the observed increase, including the role of non-COVID overall mortality. All-cause mortality data is a critical data source describing the population impact of COVID-19 in Canada, and this study has demonstrated the utility of cremation data to provide more timely mortality information during a public health emergency. Sustained monitoring of mortality trends from all data resources throughout the pandemic, including subsequent waves, is needed.

## Data Availability

A data base containing cremation certificates is held at the Office of Chief Coroner for Ontario. The data-sharing agreement for this study prohibits the data from being publicly available. Data requests may be granted provided there is an appropriate data-sharing agreement in place.

## Acknowledgements

The authors acknowledge Dr. Dirk Huyer, Chief Coroner for Ontario for the vision and support to analyze cremation certificates as a timely source for mortality data.

## Contributors

RM developed the idea for the study and coordinated the collaboration. GP conducted all analysis and RP, MD had access to the data. All authors contributed to the conception and design of the work; acquisition, analysis and interpretation of data; drafting the work and revising it critically. All authors gave final approval to the version to be published and agreed to be held accountable for all aspects of the work.

## Data sharing

A database containing cremation certificates is held at the Office of Chief Coroner for Ontario. The data-sharing agreement for this study prohibits the data from being publicly available. Data requests may be granted, provided there is an appropriate data-sharing agreement in place.

## Ethics

This study was approved by the Research Ethics Board of Western University. (Project ID: 112478)

